# Nephron Number and Kidney Outcomes in IgA Nephropathy: A Retrospective Cohort Study

**DOI:** 10.1101/2025.05.11.25327402

**Authors:** Hirokazu Marumoto, Takaya Sasaki, Nobuo Tsuboi, Vivette D. D’Agati, Yusuke Okabayashi, Kotaro Haruhara, Go Kanzaki, Kentaro Koike, John F. Bertram, Toshiharu Ninomiya, Takashi Yokoo.

## Abstract

**Background:** We previously reported substantial variability in the number of nephrons in patients with IgA nephropathy (IgAN), even among patients with similar risk factor profiles. This retrospective cohort study aimed to evaluate the clinical significance of nephron number at diagnostic biopsy for subsequent kidney outcomes in patients with IgAN.

**Methods:** The number of nephrons, defined as the total number of non-globally sclerotic glomeruli per kidney, was estimated using computed tomography imaging and biopsy-based stereology. Kidney outcomes were compared based on tertiles of nephron number. The primary endpoint was the annual slope of the estimated glomerular filtration rate (eGFR), and the secondary endpoint was the initiation of kidney replacement therapy.

**Results:** A total of 222 Japanese adults with IgAN were included. Among the entire cohort, eGFR exhibited a gradual decline over time during a median follow-up of 7.6 years. Annual eGFR slopes, adjusted for baseline eGFR, baseline proteinuria, Oxford classification scores, and therapies during the first year after biopsy, were -1.35, -1.11, and -0.97 mL/min/1.73 m^2^/year for the lowest to highest nephron number tertiles, respectively (P for trend < 0.001). Kidney replacement therapy was initiated in 32.4%, 10.8%, and 0% of patients in the lowest, middle, and highest tertiles, respectively (P for trend < 0.001). A significantly higher risk of kidney replacement therapy initiation with decreasing number of nephrons was confirmed using age- and sex-adjusted Cox proportional hazards models.

**Conclusion:** The total number of non-globally sclerotic glomeruli per kidney identified at diagnostic biopsy was independently and inversely associated with the rate of future kidney function decline in IgAN patients. As a readily quantifiable marker, it offers additional information beyond conventional clinical and histopathological risk factors. Incorporating this metric into routine evaluation may enhance risk stratification and inform more personalized, targeted treatment strategies for patients with IgAN.

**Key points:**

- No prior studies have examined the clinical significance of nephron number in patients with IgA nephropathy (IgAN).
- This study provides the first robust evidence that the number of non-globally sclerotic glomeruli per kidney is significantly and inversely associated with the rate of subsequent decline in kidney function in patients with IgAN, independent of established risk factors.
- These findings highlight the potential utility of incorporating this metric into diagnostic evaluations to help assess risk and tailor treatment for IgAN.

## Introduction

IgA nephropathy (IgAN) is the most common form of primary chronic glomerulonephritis worldwide and remains a leading cause of progressive kidney disease[1]. Approximately 30–40% of patients with IgAN progress to end-stage kidney disease within 10 to 25 years following their diagnostic biopsy[2, 3]. This progression often occurs despite treatment, underscoring the importance of identifying reliable markers that reflect disease status and therapeutic effects.

A research group at the Mayo Clinic developed a novel method that combines computed tomography imaging with biopsy-based stereology, enabling the in vivo estimation of the number of nephrons per kidney in living patients[4]. In our previous study, we used this method to estimate the number of nephrons in patients with IgAN, and in a prior cross-sectional analysis, we evaluated the association between nephron number and clinicopathological risk factors related to IgAN progression[5]. Notably, we observed substantial inter-individual variability in the number of nephrons, even among patients with similar clinical risk profiles. This finding suggested that nephron number may serve as a more fundamental indicator of kidney functional reserve beyond conventional clinical and histopathological parameters.

Building upon these findings, the present retrospective cohort study aims to determine whether nephron number at the time of diagnostic biopsy is associated with future kidney outcomes in patients with IgAN. Specifically, we seek to evaluate whether this metric serves as an independent factor associated with IgAN progression after accounting for established clinical and histopathological risk factors.

## Methods

### Patient Selection

This study included adult patients (≥18 years) who underwent native kidney biopsy at Jikei Hospital, Tokyo, between 2007 and 2017, were histopathologically diagnosed with primary IgAN, and had a minimum follow-up duration of 12 months. The diagnosis of IgAN was established based on characteristic biopsy findings of glomerulonephritis with focal or diffuse mesangial proliferative features and dominant or co-dominant deposits of IgA. Glomerular IgA deposition was confirmed by immunohistochemistry or immunofluorescence, and electron microscopy was used to identify characteristic electron dense deposits in the mesangial and para-mesangial regions. Patients with systemic diseases known to be associated with glomerular IgA deposition, such as IgA vasculitis, liver cirrhosis, systemic lupus erythematosus, and lymphoproliferative disorders, were excluded. At Jikei Hospital, kidney morphology is routinely assessed using unenhanced computed tomography imaging before performing percutaneous kidney biopsies unless there are contraindications. For this study, the exclusion criteria were defined as: (i) patients for whom computed tomography images taken within one year prior to kidney biopsy were unavailable; and (ii) patients whose kidney biopsy specimens contained fewer than five glomeruli (excluding globally sclerotic glomeruli) on light microscopy or had a cortical area of less than 2 mm².

Since this was a retrospective cohort study, the study protocol was disclosed to the public, giving patients the opportunity to opt out. Therefore, individual informed consent was not required. The study was approved by the ethics review board of the Jikei University School of Medicine (30-385 [9406]) and conducted in accordance with the principles of the Declaration of Helsinki.

### Patient Follow-up and Data Collection

Demographic and clinical data were collected at the time of kidney biopsy. Patient data during the follow-up period were obtained at intervals not exceeding six-months. Routine blood and urine samples were collected at each visit throughout the follow-up. The observation period was defined as the duration from baseline to the last clinical visit for each patient. Clinical data up to April 30, 2020, were collected and used for analysis.

### Histopathological Analysis

Kidney tissue specimens were obtained through percutaneous needle biopsy, embedded in paraffin, and sectioned at a thickness of approximately 3 µm. The sections were stained using hematoxylin-eosin, periodic acid-Schiff, Masson’s trichrome, and periodic acid silver-methenamine stains. All biopsy samples underwent immunohistochemical or immunofluorescence staining with a standard antibody panel targeting IgG, IgA, IgM, C3, and C1q. Nephron number per kidney was estimated from the number of non-globally sclerotic glomeruli identified in the kidney biopsy specimen. For this purpose, glomeruli exhibiting segmental glomerulosclerosis or extracapillary hypercellularity (crescents) were also classified as non-globally sclerotic glomeruli. The Oxford scores for mesangial hypercellularity (M), endocapillary hypercellularity (E), segmental sclerosis or adhesion (S), interstitial fibrosis and tubular atrophy (T) and crescents (C) were determined as described previously[6, 7]. Biopsy chronicity scores were calculated based on the percentages of glomerulosclerosis, interstitial fibrosis and tubular atrophy, and the severity of arteriosclerosis, as previously described[8].

### Morphological Measurements

Computed tomography images were acquired with a slice thickness of 5.0 mm. Kidney parenchymal volumes were measured using ITK-SNAP software (version 3.6, University of Pennsylvania, Philadelphia, PA, www.itksnap.org), which semi-automatically segmented the parenchymal regions from unenhanced computed tomography images of both kidneys, as previously described[9]. Kidney cortical volume was estimated using the following equation: Estimated cortical volume (cm³) = -1.3 (intercept) + 0.71 × parenchymal volume[10] (cm³). Kidney biopsy specimens were analyzed semi-automatically to measure the individual areas of all glomerular capillary tufts and the total area of the obtained kidney cortex using image analysis software (WinRoof 2017, Mitani Corporation, Tokyo, Japan)[11]. The glomerular area was defined as the mean area enclosed by the outer capillary loops of the tuft. Mean glomerular volume was calculated from the measured glomerular area as follows: 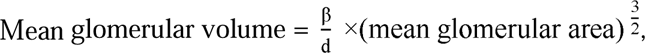 where β is a dimensionless shape coefficient (β = 1.382), and d is a size distribution coefficient (d = 1.01)[12]. The volumetric glomerular density excluding globally sclerotic glomeruli (number per mm^3^ of the cortex in biopsy) was determined based on the Weibel and Gomez stereological method as follows: 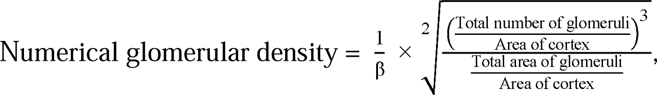 where β is a dimensionless shape coefficient (β = 1.382)[4]. The total number of nephrons was calculated by multiplying the estimated cortical volume (mm³) by the non-globally sclerotic glomerular density in both kidneys. The calculated value was divided by 2 for per kidney, by 1.43 for correcting tissue volume shrinkage due to paraffin embedding, and by 1.268 for correcting volume shrinkage due to loss of tissue perfusion pressure[4].

### Definitions

Hypertension was defined as a systolic blood pressure ≥140 mmHg, diastolic blood pressure ≥90 mmHg, or the use of antihypertensive medications. Therapies initiated within one year after diagnostic kidney biopsy, including those involving renin-angiotensin aldosterone system (RAAS) inhibitors and corticosteroids, were recorded as initial treatments. The estimated glomerular filtration rate (eGFR) was calculated using a modified equation for Japanese individuals: eGFR = 194 × age□□.²□□ × (serum creatinine)□¹.□□□ (×0.739 if female)[13]. Urinary protein excretion was assessed using either a 24-hour urine collection or a spot urine protein-to-creatinine ratio. The red blood cell (RBC) count in urinary sediment was graded as follows: grade 0, <5/high power field (HPF); grade 1, 5–9/HPF; grade 2, 10–19/HPF; grade 3, 20–49/HPF; grade 4, 50–99/HPF; and grade 5, >99/HPF.

For comparisons of kidney function outcomes, patients were divided into tertile groups based on the total number of nephrons. The primary endpoint was defined as the annual eGFR slope (mL/min/1.73 m^2^/year) calculated by a linear mixed-effect model for repeated measures. The secondary endpoint was defined as the initiation of kidney replacement therapy. The initiation of kidney replacement therapy was defined as the date on which maintenance dialysis was started or the date of kidney transplantation.

### Statistical Analysis

Baseline patient characteristics were presented as mean ± standard deviation or median [interquartile range] for continuous variables, and as frequencies and proportions for categorical variables. The Jonckheere-Terpstra test, Cochran-Armitage test, and linear regression models were used to assess trends in baseline characteristics and morphological measurements across tertile groups stratified by nephron number.

For the primary analysis, a linear mixed-effect model for repeated measures was employed to estimate the annual eGFR slope by modeling the relationship between follow-up time and eGFR levels. An unstructured covariance matrix was used for the random intercepts to account for individual variability, while a first-order autoregressive covariance structure was used to model within-subject correlation across repeated measurements. The effect of nephron number on the eGFR slope was evaluated by incorporating an interaction term between nephron number and follow-up time into the model. The assumption of linearity between follow-up time and eGFR was assessed visually using a generalized additive model, which applied data smoothing to examine the underlying trend.

A subgroup analysis was conducted based on categories of known clinical and histopathological risk factors for the progression of IgAN. These included baseline eGFR, baseline proteinuria, the Oxford classification scores, use of RAAS inhibitors within one year after diagnosis, and use of corticosteroids within one year after diagnosis. Sensitivity analyses were performed in patients whose number of nephrons fell within the 5th to 95th percentiles, those with biopsy specimens containing a cortical area greater than 4 mm², and those with a follow-up duration of 24 months or longer.

For the secondary endpoint, the cumulative incidence of kidney replacement therapy initiation was estimated using Kaplan-Meier survival curves, and comparisons were made using the log-rank test. Survival time for each patient was defined as the duration from biopsy diagnosis to the final follow-up visit. Cox proportional hazards models were used to estimate hazard ratios for kidney replacement therapy initiation.

A two-sided P-value <0.05 was considered statistically significant. To account for multiple comparisons in the subgroup analysis, Bonferroni correction was applied to the P for interaction to avoid false positives. All statistical analyses, except for generalized additive model fitting, were performed using SAS version 9.4 (SAS Institute Inc., Cary, NC, USA). The generalized additive model was analyzed using R version 4.2.2 (R Foundation for Statistical Computing, Vienna, Austria).

## Results

### Patient characteristics at baseline and during follow-up

The patient selection process is shown in **Supplemental Figure S1**. A total of 324 patients diagnosed with IgAN at a single institution between 2007 and 2017 were retrospectively identified. Among them, 222 patients were included in the present study based on inclusion and exclusion criteria. The characteristics of all patients and those of tertile groups based on nephron number are shown in **Table 1**. Overall, the mean age at diagnostic biopsy was 42.7 years, and 137 patients (61.7%) were male. The mean eGFR was 60.6 mL/min/1.73m^2^, and the median proteinuria was 814 mg/day. With decreasing nephrons, patients were older, more likely to be hypertensive, more likely to be treated with pre-biopsy RAAS inhibitors and had kidney dysfunction and heavy proteinuria. Regardless of nephron number, there were no differences between groups in the total number of glomeruli obtained from biopsy specimens, the proportion of globally sclerotic glomeruli, or the chronicity score. Scores for Oxford classification were not different between tertile groups. For the entire IgAN cohort, the mean number of nephrons per kidney was 680,000 ± 398,000. The low, middle, and high tertile subgroups had averages of 280,000±109,000, 628,000±104,000, and 1,131,000±292,000 nephrons, respectively. With regard to treatment within the first year after biopsy, patients with lower nephron numbers were treated more frequently with RAAS inhibitors than those with higher nephron numbers, while treatment with corticosteroids did not differ between the three groups.

**Table 1.**
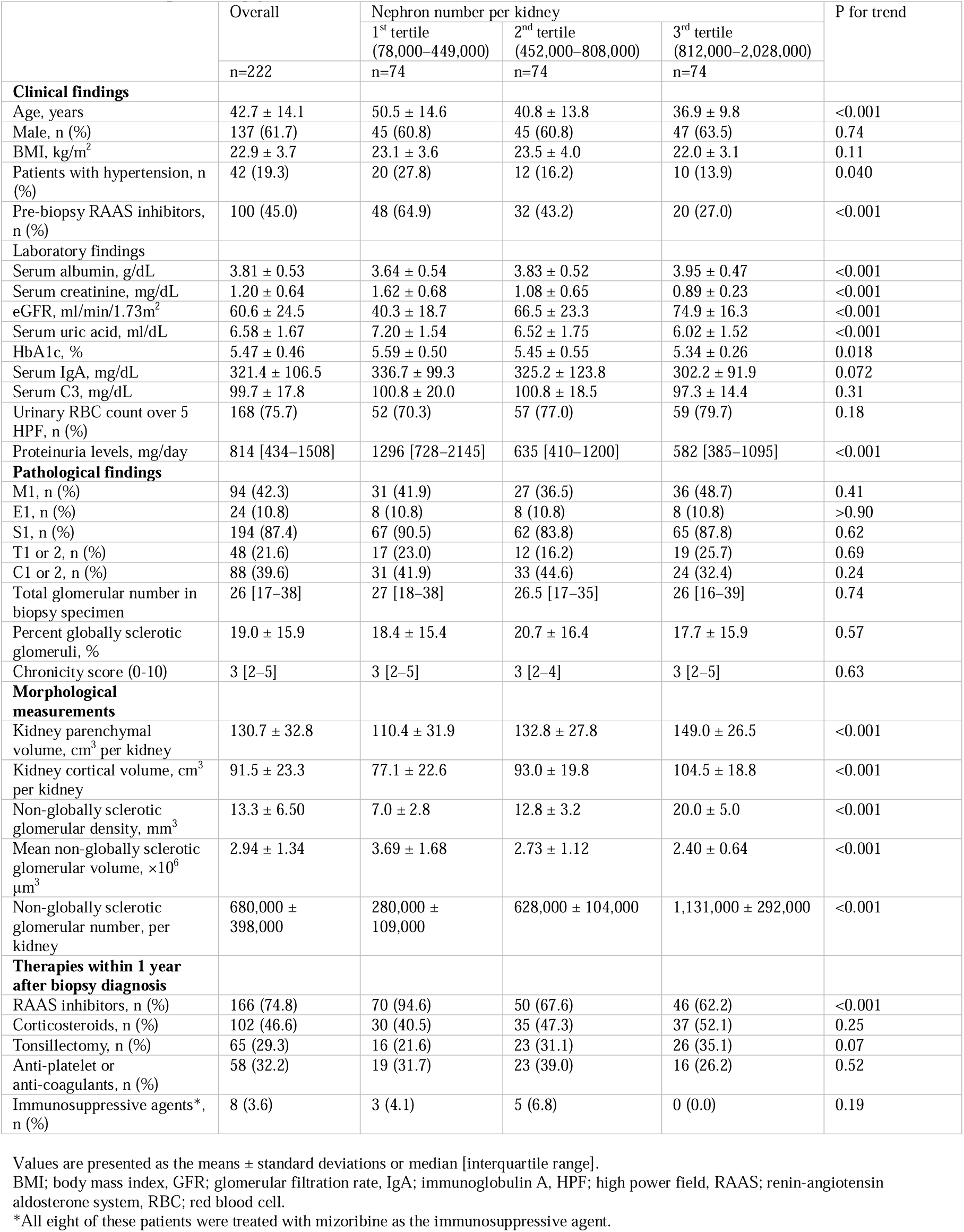
Clinicopathological and morphometric findings, and treatments during the first year in all patients and by nephron number tertiles at diagnostic biopsy.

|  | Overall | Nephron number per kidney |  |  | P for trend |
| --- | --- | --- | --- | --- | --- |
|  |  | 1<sup>st</sup> tertile<br>(78,000–449,000) | 2<sup>nd</sup> tertile<br>(452,000–808,000) | 3<sup>rd</sup> tertile<br>(812,000–2,028,000) |  |
|  | n=222 | n=74 | n=74 | n=74 |  |
| **Clinical findings** |  |  |  |  |  |
| Age, years | 42.7 ± 14.1 | 50.5 ± 14.6 | 40.8 ± 13.8 | 36.9 ± 9.8 | <0.001 |
| Male, n (%) | 137 (61.7) | 45 (60.8) | 45 (60.8) | 47 (63.5) | 0.74 |
| BMI, kg/m<sup>2</sup> | 22.9 ± 3.7 | 23.1 ± 3.6 | 23.5 ± 4.0 | 22.0 ± 3.1 | 0.11 |
| Patients with hypertension, n (%) | 42 (19.3) | 20 (27.8) | 12 (16.2) | 10 (13.9) | 0.040 |
| Pre-biopsy RAAS inhibitors, n (%) | 100 (45.0) | 48 (64.9) | 32 (43.2) | 20 (27.0) | <0.001 |
| **Laboratory findings** |  |  |  |  |  |
| Serum albumin, g/dL | 3.81 ± 0.53 | 3.64 ± 0.54 | 3.83 ± 0.52 | 3.95 ± 0.47 | <0.001 |
| Serum creatinine, mg/dL | 1.20 ± 0.64 | 1.62 ± 0.68 | 1.08 ± 0.65 | 0.89 ± 0.23 | <0.001 |
| eGFR, ml/min/1.73m<sup>2</sup> | 60.6 ± 24.5 | 40.3 ± 18.7 | 66.5 ± 23.3 | 74.9 ± 16.3 | <0.001 |
| Serum uric acid, ml/dL | 6.58 ± 1.67 | 7.20 ± 1.54 | 6.52 ± 1.75 | 6.02 ± 1.52 | <0.001 |
| HbA1c, % | 5.47 ± 0.46 | 5.59 ± 0.50 | 5.45 ± 0.55 | 5.34 ± 0.26 | 0.018 |
| Serum IgA, mg/dL | 321.4 ± 106.5 | 336.7 ± 99.3 | 325.2 ± 123.8 | 302.2 ± 91.9 | 0.072 |
| Serum C3, mg/dL | 99.7 ± 17.8 | 100.8 ± 20.0 | 100.8 ± 18.5 | 97.3 ± 14.4 | 0.31 |
| Urinary RBC count over 5 HPF, n (%) | 168 (75.7) | 52 (70.3) | 57 (77.0) | 59 (79.7) | 0.18 |
| Proteinuria levels, mg/day | 814 [434–1508] | 1296 [728–2145] | 635 [410–1200] | 582 [385–1095] | <0.001 |
| **Pathological findings** |  |  |  |  |  |
| M1, n (%) | 94 (42.3) | 31 (41.9) | 27 (36.5) | 36 (48.7) | 0.41 |
| E1, n (%) | 24 (10.8) | 8 (10.8) | 8 (10.8) | 8 (10.8) | >0.90 |
| S1, n (%) | 194 (87.4) | 67 (90.5) | 62 (83.8) | 65 (87.8) | 0.62 |
| T1 or 2, n (%) | 48 (21.6) | 17 (23.0) | 12 (16.2) | 19 (25.7) | 0.69 |
| C1 or 2, n (%) | 88 (39.6) | 31 (41.9) | 33 (44.6) | 24 (32.4) | 0.24 |
| Total glomerular number in biopsy specimen | 26 [17–38] | 27 [18–38] | 26.5 [17–35] | 26 [16–39] | 0.74 |
| Percent globally sclerotic glomeruli, % | 19.0 ± 15.9 | 18.4 ± 15.4 | 20.7 ± 16.4 | 17.7 ± 15.9 | 0.57 |
| Chronicity score (0-10) | 3 [2–5] | 3 [2–5] | 3 [2–4] | 3 [2–5] | 0.63 |
| **Morphological measurements** |  |  |  |  |  |
| Kidney parenchymal volume, cm<sup>3</sup> per kidney | 130.7 ± 32.8 | 110.4 ± 31.9 | 132.8 ± 27.8 | 149.0 ± 26.5 | <0.001 |
| Kidney cortical volume, cm<sup>3</sup> per kidney | 91.5 ± 23.3 | 77.1 ± 22.6 | 93.0 ± 19.8 | 104.5 ± 18.8 | <0.001 |
| Non-globally sclerotic glomerular density, mm<sup>3</sup> | 13.3 ± 6.50 | 7.0 ± 2.8 | 12.8 ± 3.2 | 20.0 ± 5.0 | <0.001 |
| Mean non-globally sclerotic glomerular volume, ×10<sup>6</sup> μm<sup>3</sup> | 2.94 ± 1.34 | 3.69 ± 1.68 | 2.73 ± 1.12 | 2.40 ± 0.64 | <0.001 |
| Non-globally sclerotic glomerular number, per kidney | 680,000 ± 398,000 | 280,000 ± 109,000 | 628,000 ± 104,000 | 1,131,000 ± 292,000 | <0.001 |
| **Therapies within 1 year after biopsy diagnosis** |  |  |  |  |  |
| RAAS inhibitors, n (%) | 166 (74.8) | 70 (94.6) | 50 (67.6) | 46 (62.2) | <0.001 |
| Corticosteroids, n (%) | 102 (46.6) | 30 (40.5) | 35 (47.3) | 37 (52.1) | 0.25 |
| Tonsillectomy, n (%) | 65 (29.3) | 16 (21.6) | 23 (31.1) | 26 (35.1) | 0.07 |
| Anti-platelet or anti-coagulants, n (%) | 58 (32.2) | 19 (31.7) | 23 (39.0) | 16 (26.2) | 0.52 |
| Immunosuppressive agents*, n (%) | 8 (3.6) | 3 (4.1) | 5 (6.8) | 0 (0.0) | 0.19 |

Values are presented as the means ± standard deviations or median [interquartile range].
BMI; body mass index, GFR; glomerular filtration rate, IgA; immunoglobulin A, HPF; high power field, RAAS; renin-angiotensin aldosterone system, RBC; red blood cell.
\*All eight of these patients were treated with mizoribine as the immunosuppressive agent.

### Annual estimated glomerular filtration rate slope

Overall, the mean follow-up duration was 7.56 ± 3.86 years, with no significant difference between the nephron number tertiles: 7.02 ± 3.91 years in the first tertile, 7.75 ± 3.71 years in the second, and 7.91 ± 3.94 years in the third. **Figure 1A** shows a spaghetti plot of eGFR during follow-up for each patient in time series. In this cohort of patients with IgAN, kidney functional decline was generally slowly progressive. **Figure 1B** illustrates the annual eGFR slope for each group based on nephron number using a linear mixed-effect model for repeated measures. The model was adjusted for age, sex, eGFR at baseline, log-transformed proteinuria at baseline, Oxford classification scores (i.e., M, E, S, T, and C score), use of RAAS inhibitors during the first year, and use of corticosteroids during the first year. The annual decline in eGFR was fastest in patients within the lowest tertile of nephrons (tertile 1), with a slope of –1.35 mL/min/1.73 m²/year (95% CI: –1.41 to –1.29, P <0.001). By comparison, the slope was –1.11 mL/min/1.73 m²/year (95% CI: –1.18 to –1.03, P <0.001) in tertile 2 and –0.97 mL/min/1.73 m²/year (95% CI: –1.05 to –0.89, P <0.001) in tertile 3, indicating a slower rate of eGFR decline in patients with a higher nephron count at the commencement of the study. The difference in eGFR slope between tertile 1 and tertile 3 was statistically significant (–0.34 mL/min/1.73 m²/year, 95% CI: –0.55 to –0.13, P = 0.001), while the difference between tertile 2 and tertile 3 did not reach statistical significance (–0.18 mL/min/1.73 m²/year, 95% CI: –0.40 to +0.04, P = 0.12). Overall, a significant trend was observed, showing that a higher number of nephrons was associated with a slower decline in eGFR (P for trend < 0.001).

**Figure 1.**
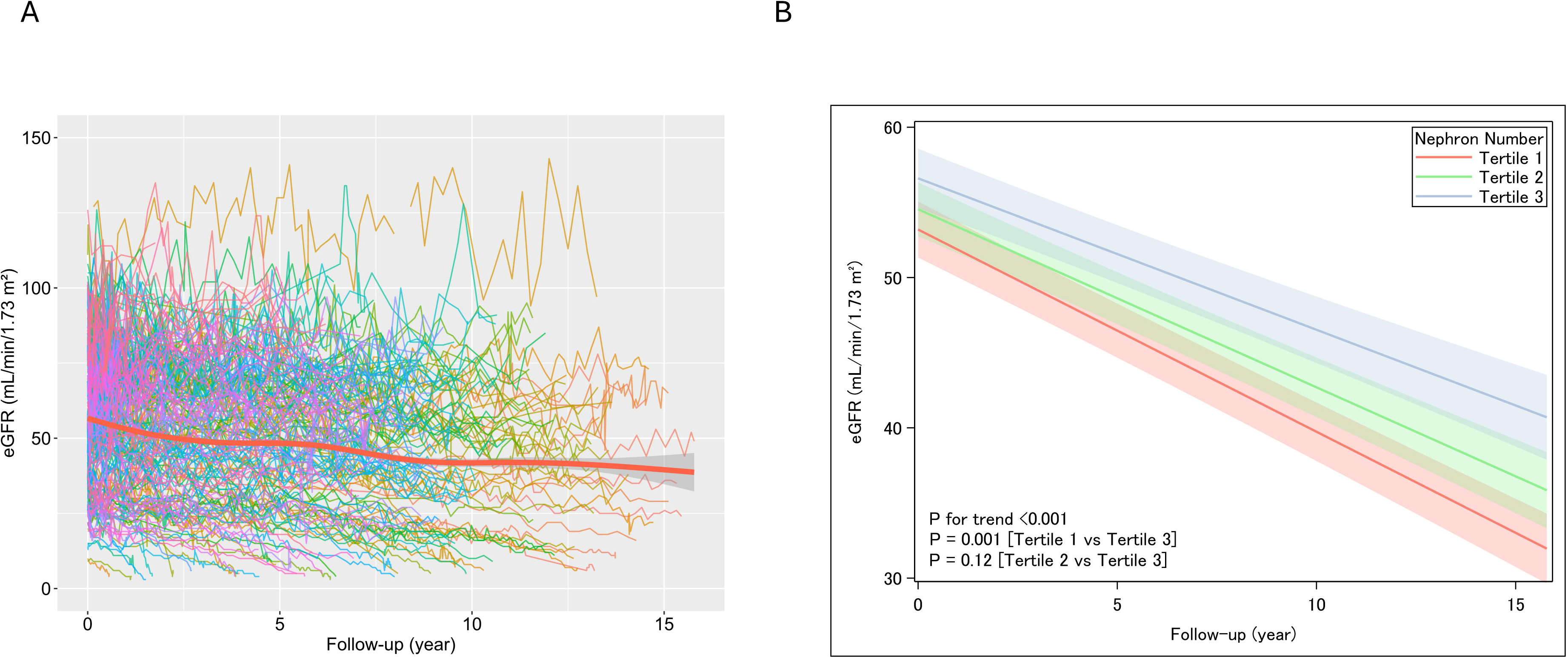
Longitudinal changes in estimated glomerular filtration rate in individual cases and intergroup comparisons according to nephron number. A spaghetti plot of eGFR over the follow-up period for each patient is shown in time series, with the bold line indicating the overall mean value (**A**). A mixed model for repeated measures was used to model the association between eGFR and follow-up time, and calculate the slope and P value for each group (**B**). The model was adjusted for age, sex, eGFR at baseline, log-transformed proteinuria at baseline, the Oxford pathological classification scores (i.e., M, E, S, T, and C score), use of renin-angiotensin-aldosterone inhibitors during the first year, and use of corticosteroids during the first year. eGFR, estimated glomerular filtration rate

### Subgroup and sensitivity analyses

Subgroup analyses showed that most subgroups stratified by clinical risk factors had point estimates that fell within the 95% confidence intervals of the overall population, suggesting consistent associations across different patient profiles, despite significant interactions in subgroups defined by baseline eGFR, the M and S scores of the Oxford classification (**Figure 2**). Sensitivity analyses restricting the study population did not substantially alter the results (**Supplemental Figure S2**).

**Figure 2.**
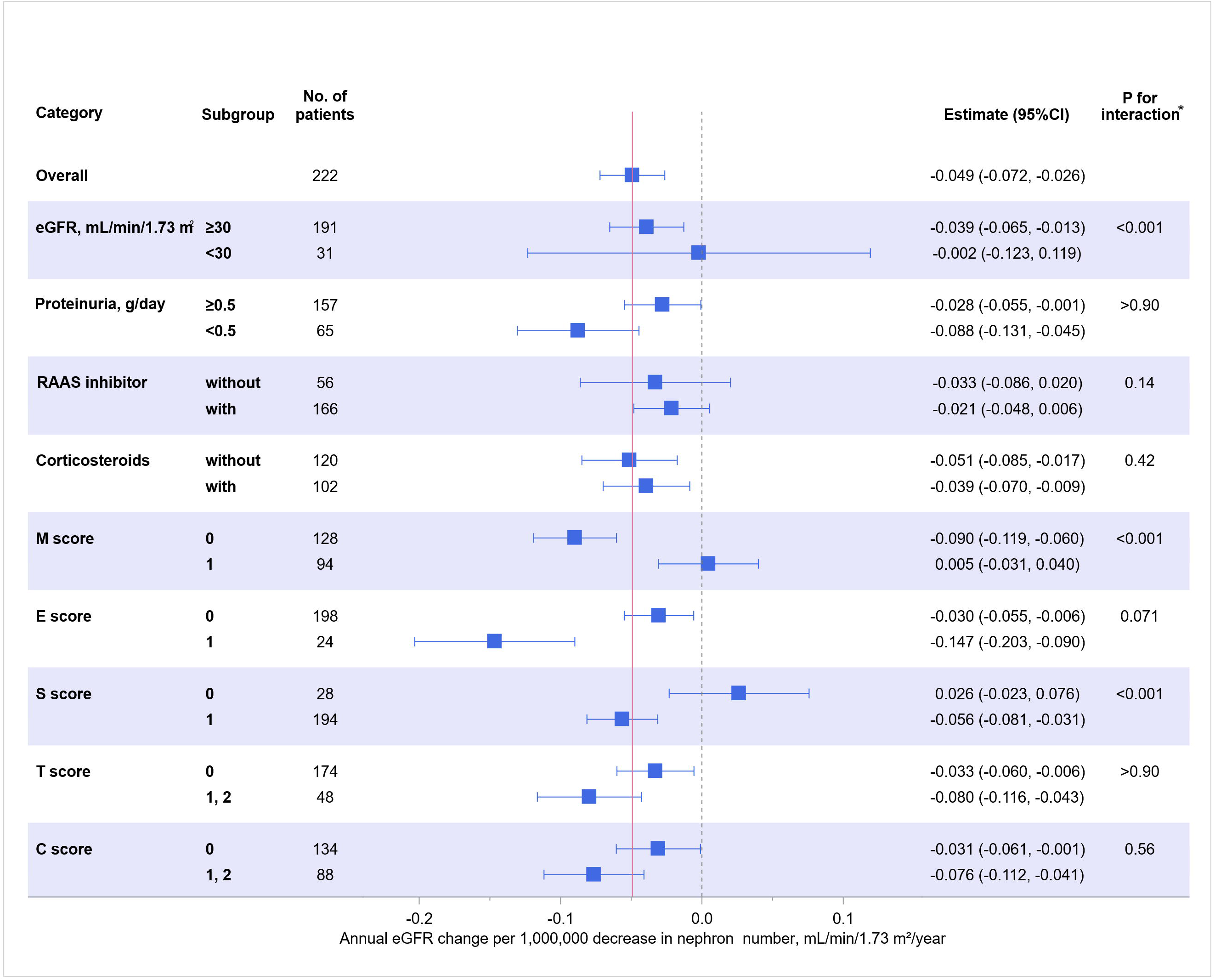
Subgroup analyses for annual estimated glomerular filtration rate slope as per decrease in nephron number. The estimates of annual eGFR slope (mL/min/1.73m²/year) with 95% confidence intervals (CIs) per 10^6^ decrease in nephrons, stratified by baseline eGFR, and baseline proteinuria, the Oxford pathological classification scores (i.e., M, E, S, T, and C score), RAAS inhibitor treatment within one year after diagnosis, and corticosteroid treatment within one year after diagnosis are presented. P-values were adjusted using the Bonferroni correction. The pink solid vertical line indicates the eGFR slope estimate for the overall patients group. eGFR, estimated glomerular filtration rate; RAAS, renin-angiotensin aldosterone system

### Incidence of kidney replacement therapy initiation

During the follow-up period, 32 of all 222 patients (14.3%) initiated kidney replacement therapy. Based on nephron number tertiles, 24 patients (32.4%) in tertile 1, 8 patients (10.8%) in tertile 2, and none in tertile 3 underwent kidney replacement therapy initiation. The cumulative incidence of kidney replacement therapy initiation was significantly higher in patients with fewer nephrons (P for trend <0.001, **Figure 3**). Log-rank testing confirmed a significant difference in the incidence of kidney replacement therapy across nephron tertiles. Furthermore, both crude and age- and sex-adjusted Cox proportional hazards models revealed a sequential increase in the hazard ratio for kidney replacement therapy initiation with decreasing nephron number (**Table 2**).

**Figure 3.**
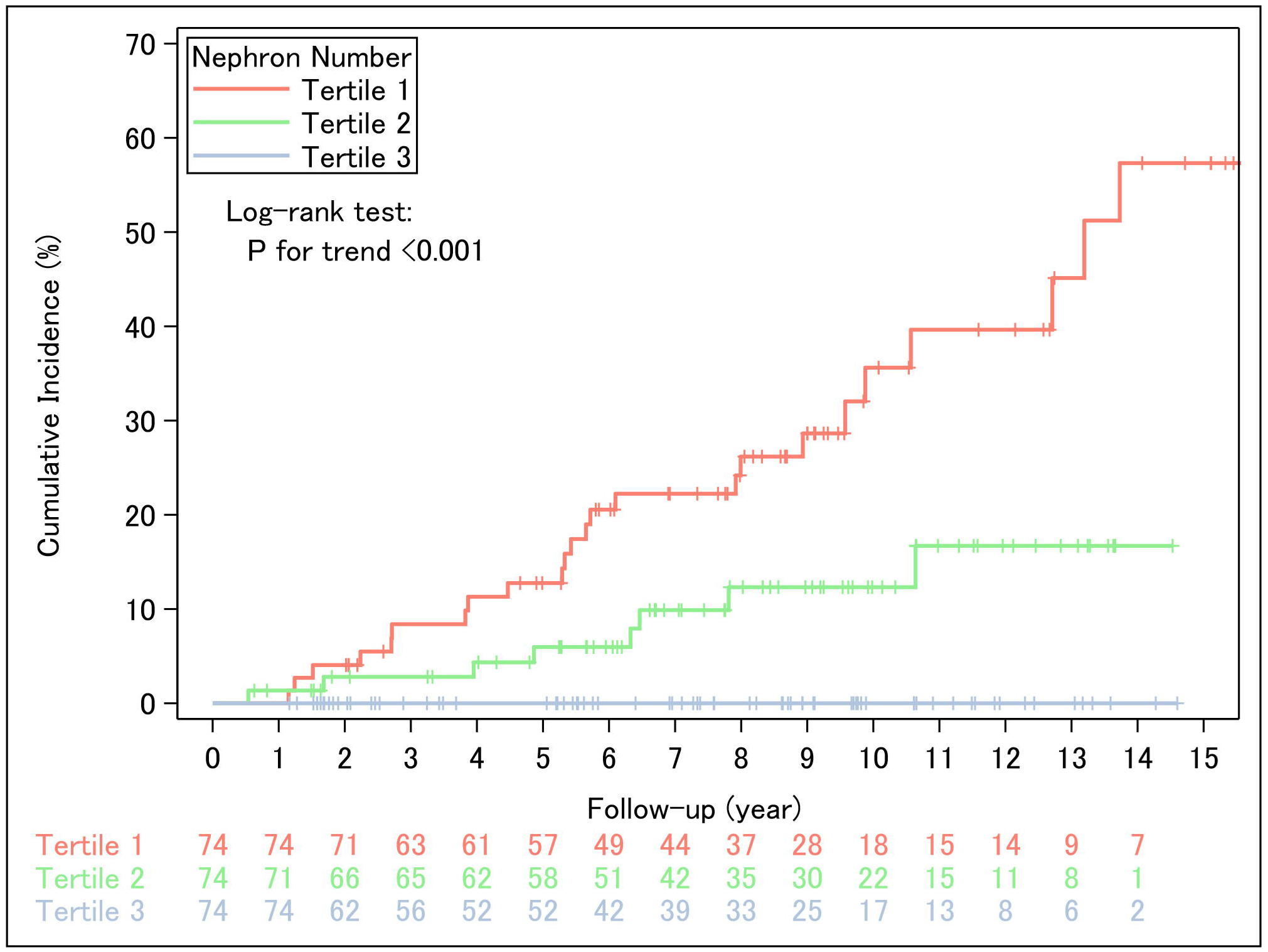
Cumulative incidence for kidney replacement therapy initiation based on nephron number. The cumulative incidence for kidney replacement therapy initiation, stratified by tertiles of nephron number, is presented using an inverse Kaplan-Meier curve. The P for trend was calculated using the log-rank test for trend.

**Table 2.** Hazard ratio of nephron number for the initiation of kidney replacement therapy.

|  | Nephron number<br>(per 100,000 decrease / kidney) |  |  |
| --- | --- | --- | --- |
|  | Hazard ratio | (95% confidence interval) | P value |
| Crude | 1.47 | (1.25–1.73) | <0.001 |
| Age- and sex-adjusted | 1.42 | (1.18–1.71) | <0.001 |

Crude and age- and sex-adjusted Cox proportional hazards models analyzing the number of nephrons as an explanatory variable for the initiation of kidney replacement therapy.

## Discussion

Over the past several decades, extensive research has been conducted to elucidate the natural history of IgAN and identify the factors associated with disease progression[14-16]. While numerous clinical and histopathological markers have been proposed for risk stratification, nephron number has received relatively little attention in this context. In this study, we estimated the number of nephrons per kidney and analyzed their association with IgAN progression. We found that nephron number was independently associated with the eGFR slope during follow-up, a marker increasingly recognized for its ability to capture disease progression and treatment response over time in patients with chronic kidney disease, including IgAN[17-19]. This slope was estimated using a linear mixed-effects model for repeated measures, which enabled individualized and robust analysis of longitudinal data without the need for imputation. The association remained significant even after adjusting for established clinical and histopathological risk factors. Furthermore, similar findings were observed when using kidney replacement therapy initiation as a definitive outcome, underscoring the clinical significance of nephron number in IgAN.

Human nephron number shows wide inter-individual variation at birth and decreases over time, primarily due to glomerulosclerosis that occurs with aging[4, 20, 21]. In the presence of kidney disease, this age-related decline may be further exacerbated by ongoing pathological processes[22]. In IgAN, most patients remain asymptomatic until later stages, except for those with gross hematuria. Consequently, the timing of nephrology referral and therapeutic intervention varies considerably among individuals, leading to significant heterogeneity in clinical presentation and disease severity at diagnosis[23, 24]. Thus, nephron number at diagnostic biopsy reflects a complex interplay between congenital endowment, age-related loss, and disease-driven injury and scarring. Our findings therefore demonstrate that nephron number at diagnostic biopsy offers additional information beyond conventional markers, highlighting its potential role in refining risk stratification and guiding individualized therapeutic strategies in patients with IgAN.

In this study, patients in the lowest nephron tertile exhibited not only fewer nephrons per kidney but also significantly larger mean glomerular volumes compared to those in the highest tertile, consistent with compensatory hypertrophy at the single-nephron level. These patients also demonstrated lower baseline eGFR and greater proteinuria at the time of biopsy, suggesting that nephron loss had already progressed by the time of diagnosis. Despite the similar total number of glomeruli sampled at biopsy and the similar proportion of globally sclerotic glomeruli across the three groups, the larger glomerular cross-sectional area observed in the group with fewest nephrons appears to be the primary factor contributing to the lower estimated glomerular density. In addition, previously existing glomeruli may have been selectively lost by resorption into the surrounding tissue[4, 25], contributing to overall cortical atrophy and a reduction in total number of nephrons. Consequently, morphometric indices that incorporate cortical volume and non-sclerotic glomerular density, as employed in this study, may offer more accurate and robust estimates of nephron loss than relying solely on the proportion of globally or non-globally sclerotic glomeruli observed in biopsy specimens as assessed in routine histopathological diagnosis.

IgAN is characterized by a broad spectrum of glomerular lesions; therefore, the relevance of nephron number should be evaluated in the context of disease-specific structural alterations. Although variability and interaction observed in the subgroup analyses of the present study suggest that the strength of association may vary depending on histopathological factors such as the presence of Oxford classification lesions, these findings should be interpreted with caution due to the limited sample sizes and the exploratory nature of the analysis. Since the number of nephrons provides information about kidney structural reserve that complements existing histopathological indices, it may offer additional perspectives relevant to treatment planning for IgAN [26]. Nephron number may serve as a robust anatomical reference point, offering a valuable framework for monitoring long-term treatment effects beyond inflammatory changes. Thus, incorporating nephron number may help determine the indications for various evidence-based treatments for IgAN, including RAAS inhibitors[27] and sodium-glucose cotransporter 2 inhibitors[18]. These treatments may be more appropriately applied from an early disease stage and continued over the long term. The future application of this metric to a wider range of patient populations, encompassing diverse active and chronic histopathological lesions, is expected to enhance our understanding of the pathophysiology of IgAN.

Despite the strengths of our study, several limitations should be acknowledged. First, the relatively small sample size may have limited the statistical power to detect associations, particularly regarding definitive kidney outcomes such as the initiation of kidney replacement therapy. This limitation reduced our ability to adjust for potential confounding variables in Cox regression models and to assess the influence of kidney biopsy findings on disease progression, with or without treatment effects. Second, the retrospective design of the study introduces the possibility of selection bias in patient inclusion and treatment decisions. Although gender differences in IgAN incidence are generally minimal, our study cohort showed a slight male predominance. This may partly reflect lower use of computed tomography among women due to concerns about radiation exposure. Third, although our sensitivity analysis accounting for biopsy adequacy supports the robustness of the results, estimating nephron number from needle biopsy specimens has inherent limitations, particularly due to potential variability in glomerular distribution across the kidney[28]. Fourth, our cohort lacks data on indicators of nephron endowment, such as birth weight. Therefore, it is difficult to assess the relative contributions of nephron loss due to IgAN and low nephron endowment at birth. Finally, as this was a single-center study conducted entirely in a Japanese population, the generalizability of our findings to other ethnic or geographic populations remains uncertain. In Japan, routine urinalysis screening enables earlier detection of IgAN than in many other countries[29], which may influence the clinical and pathological features of the study cohort. Future studies involving more diverse populations are needed to confirm the applicability of these findings in broader clinical settings.

In conclusion, this study provides the first robust evidence that nephron number at the time of diagnostic biopsy is significantly and inversely associated with subsequent decline in kidney function in patients with IgAN. Given the wide variability in nephron number among individuals, integrating this parameter into clinical decision-making may improve the precision of IgAN management. Future prospective studies with larger cohorts and longer follow-up periods are warranted to validate our findings and explore potential therapeutic implications.

## Authors’ Contributions

Conceptualization: Hirokazu Marumoto, Takaya Sasaki, Nobuo Tsuboi.

Data curation: Hirokazu Marumoto.

Formal analysis: Hirokazu Marumoto, Takaya Sasaki, Nobuo Tsuboi.

Funding acquisition: Nobuo Tsuboi.

Investigation: Hirokazu Marumoto, Takaya Sasaki, Nobuo Tsuboi.

Methodology: Hirokazu Marumoto, Takaya Sasaki, Nobuo Tsuboi.

Project administration: Nobuo Tsuboi.

Supervision: Nobuo Tsuboi, Takashi Yokoo.

Validation: Hirokazu Marumoto, Takaya Sasaki.

Visualization: Hirokazu Marumoto, Takaya Sasaki, Nobuo Tsuboi.

Writing – original draft: Hirokazu Marumoto, Takaya Sasaki, Nobuo Tsuboi.

Writing – review & editing: Vivette.D. D’Agati, Yusuke Okabayashi, Kotaro Haruhara, Go Kanzaki, Kentaro Koike, John. F. Bertram, Toshiharu Ninomiya, Takashi Yokoo.

## Acknowledgments

Ethics committee of the Jikei University School of Medicine gave ethical approval for this work [30-385 (9406)].

## Data availability statement

All data produced in the present study are available upon reasonable request to the authors.

