## Supplemental materials for "Nephron Number and Kidney Outcomes in IgA Nephropathy: A Retrospective Cohort Study"

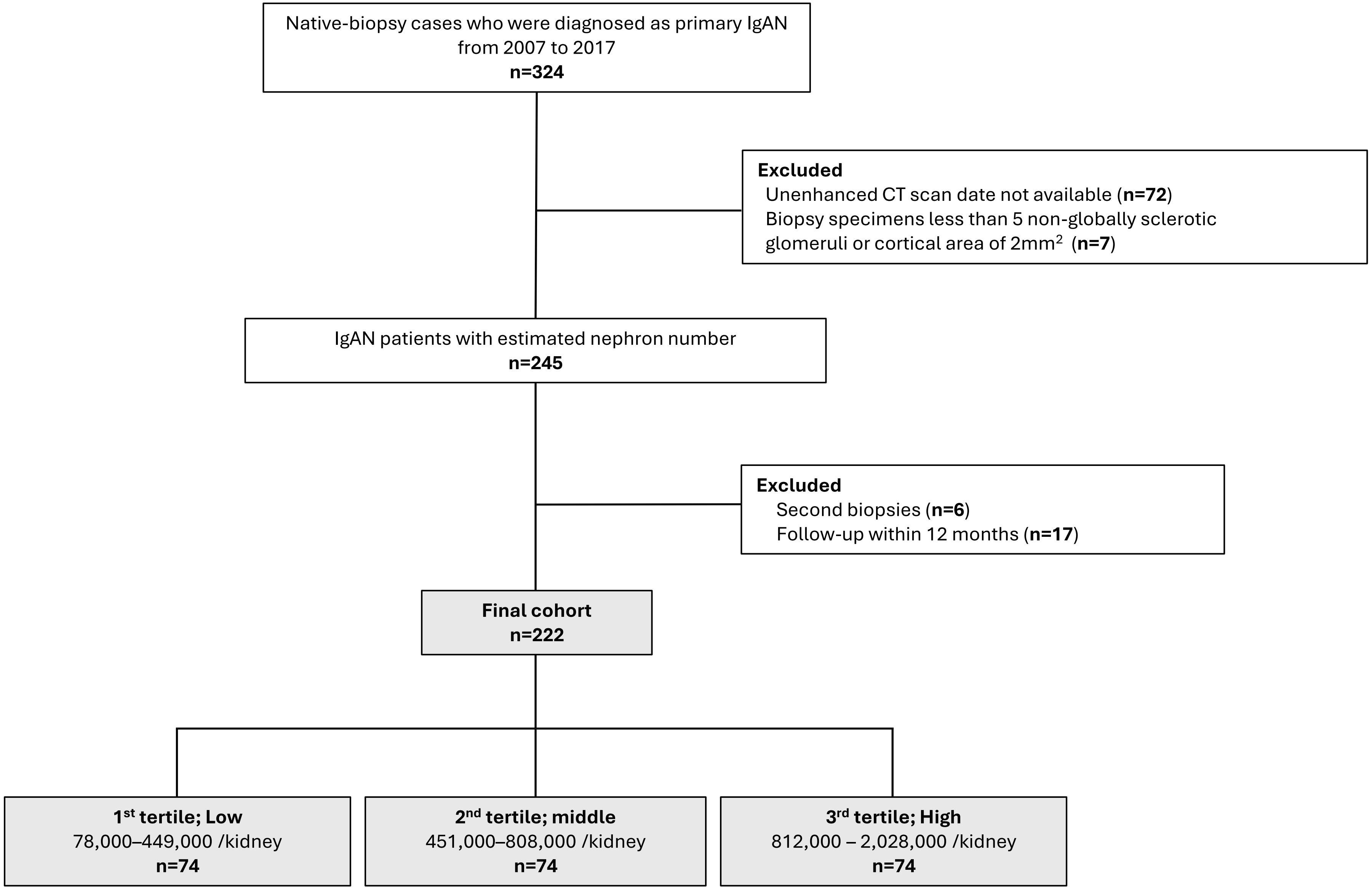


A total of 324 patients diagnosed with IgA nephropathy (IgAN) between 2007 and 2017 were retrospectively identified. Among them, 222 patients were included in the present study based on predefined inclusion and exclusion criteria.

Supplemental Figure S1.

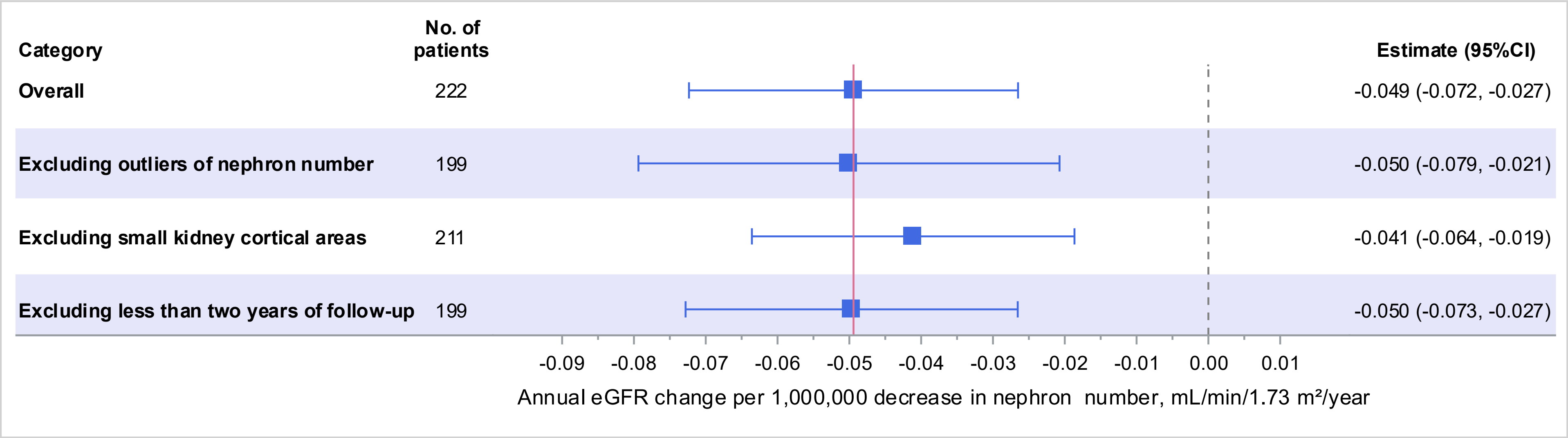


Sensitivity analyses were performed in patients whose number of nephrons within the 5th to 95th percentiles, those with biopsy specimens containing a cortical area greater than 4 $mm^2$, and those with a follow-up duration of 24 months or longer.

Supplemental Figure S2.
